# Effects of Elexacaftor/Tezacaftor/Ivacaftor on Sputum Viscoelastic Properties, Airway Infection and Inflammation in Patients with Cystic Fibrosis

**DOI:** 10.1101/2022.12.26.22283946

**Authors:** Annalisa Addante, Mirjam Völler, Laura Schaupp, Kerstin Fentker, Markus Bardua, Aditi Kuppe, Julia Duerr, Linus Piehler, Jobst Röhmel, Stephanie Thee, Marieluise Kirchner, Matthias Ziehm, Daniel Lauster, Rainer Haag, Michael Gradzielski, Mirjam Stahl, Philipp Mertins, Sébastien Boutin, Simon Y. Graeber, Marcus A. Mall

**Author notes:** **Correspondence should be addressed to:** Marcus A. Mall, M.D., Department of Pediatric Respiratory Medicine, Immunology and Critical Care Medicine, Charité - Universitätsmedizin Berlin, Augustenburger Platz 1, 13353 Berlin, Germany. A.A., M.V., L.S. and K.F. contributed equally as first authors. P.M., S.B., S.Y.G. and M.A.M. contributed equally as senior authors. **Take home message:** The triple combination CFTR modulator therapy ELX/TEZ/IVA improves viscoelastic properties of airway mucus, chronic airway infection and inflammation in CF patients with at least one *F508del* allele, however without reaching levels close to healthy.

## Abstract

**Background:** We recently demonstrated that the triple combination CFTR modulator therapy elexacaftor/tezacaftor/ivacaftor (ELX/TEZ/IVA) improves lung ventilation and airway mucus plugging determined by multiple-breath washout and magnetic resonance imaging in CF patients with at least one *F508del* allele. However, effects of ELX/TEZ/IVA on viscoelastic properties of airway mucus, chronic airway infection and inflammation have not been studied. The aim of this study was, therefore, to determine the effects of ELX/TEZ/IVA on airway mucus rheology, microbiome and inflammation in CF patients with one or two *F508del* alleles aged 12 years and older.

**Methods:** In this prospective observational study, we assessed sputum rheology, the microbiome, inflammation markers and proteome before and 8 to 16 weeks after initiation of ELX/TEZ/IVA.

**Results:** In total, 59 patients with CF and at least one *F508del* allele and 10 healthy controls were enrolled in this study. ELX/TEZ/IVA improved the elastic modulus (G’; -6.3 Pa; IQR, -17.9 to 1.2; *P*<0.01) and viscous modulus (G’’; -1.6 Pa; IQR, -3.6 to 0.5; *P*<0.05) of CF sputum. Further, ELX/TEZ/IVA improved the microbiome α-diversity (0.6; IQR, 0.0 to 1.2; *P*<0.001) and decreased the relative abundance of *Pseudomonas aeruginosa* in CF sputum. ELX/TEZ/IVA also reduced IL-8 (−11.7 ng/ml, IQR, -36.5 to 11.2; *P*<0.05) and free NE activity (−27.5 µg/ml, IQR, - 64.5 to -3.5; *P*<0.001), and shifted the CF sputum proteome towards healthy.

**Conclusions:** Our data demonstrate that ELX/TEZ/IVA improves sputum viscoelastic properties, chronic airway infection and inflammation in CF patients with at least one *F508del* allele, however, without reaching levels close to healthy.

Clinical trial registered with www.clinicaltrials.gov (NCT04732910)

## Introduction

CFTR dysfunction leads to abnormal viscoelastic properties of airway mucus, impaired mucociliary clearance and mucus plugging, which sets the stage for chronic polymicrobial infection and neutrophilic inflammation that are key drivers of the onset and progression of structural lung damage in patients with cystic fibrosis (CF) [1-6]. The triple combination CFTR modulator therapy elexacaftor/tezacaftor/ivacaftor (ELX/TEZ/IVA) showed unprecedented improvements in lung function and other clinical outcomes in pivotal clinical trials and post-approval observational studies in patients with CF with at least one copy of the common *F508del* allele [7-15]. We recently demonstrated that ELX/TEZ/IVA improves F508del-CFTR function in the airways of patients with one or two *F508del* alleles to ∼40 to 50% of normal CFTR activity [15]. This degree of restoration of CFTR function is superior to previous dual drug combinations of lumacaftor/ivacaftor (LUM/IVA) and tezacaftor/ivacaftor (TEZ/IVA) in *F508del* homozygous patients and comparable to ivacaftor monotherapy in CF patients with a *G551D* mutation [15-20]. In a previous study, we showed that this level of rescue of F508del-CFTR function achieved by ELX/TEZ/IVA leads to substantial improvement in airway mucus obstruction, as evidenced by reduced mucus plugging detected by magnetic resonance imaging and improved ventilation homogeneity detected by multiple-breath washout [14]. However, the effects of ELX/TEZ/IVA on abnormal viscoelasticity of airway mucus and chronic inflammation in patients with CF have not been studied, and data of effects on the sputum microbiome remain limited [21, 22].

The aim of this study was, therefore, to assess the effects of ELX/TEZ/IVA on sputum viscoelastic properties, chronic airway infection and inflammation in patients with CF with at least one *F508del* allele. To achieve this goal, we conducted a prospective observational study in 59 patients with CF compound-heterozygous for *F508del* and a minimal function mutation or homozygous for *F508del* and investigated rheological properties, microbiota, and key inflammation markers implicated in lung disease progression, including neutrophil elastase (NE), interleukin (IL)-1β and IL-8 [4-6], in sputum samples at baseline and 8 to 16 weeks after initiation of ELX/TEZ/IVA therapy. Further, we performed proteomics analysis as an unbiased approach to determine effects of ELX/TEZ/IVA on abnormalities of the CF sputum proteome and compared the results from patients with CF to those obtained from sputum of 10 age-matched healthy controls.

## Methods

Additional details on methods are provided in the online supplement.

### Study design and participants

This prospective observational post-approval study (clinicaltrials.gov Identifier: NCT04732910) was approved by the ethics committee of Charité - Universitätsmedizin Berlin (EA2/220/18). Written informed consent was obtained from all patients, their parents or legal guardians. Patients were eligible to participate if they were at least 12 years old, compound-heterozygous for *F508del* and a minimal function mutation or homozygous for *F508del*, and fulfilled the inclusion and exclusion criteria detailed in the online supplement. Healthy control subjects were age- and sex-matched non-smoking volunteers without any medical history of respiratory disease. Sputum rheology, microbiome analysis, cytokine measurements and proteomics were assessed if patients provided sputum at baseline and 8 to 16 weeks after initiation of therapy with the approved dose of ELX 200 mg and TEZ 100 mg every 24 hours in combination with IVA 150 mg every 12 hours (supplementary figure S1).

### Sputum collection

Sputum samples were collected after spontaneous expectoration, directly put on ice and immediately assessed for rheology and membrane-bound NE activity on sputum neutrophils, and stored at -80 °C for subsequent analyses.

### Sputum rheology

Sputum rheology was measured with a cone and plate rheometer (Kinexus Pro+, Netzsch GmbH, Selb, Germany) as previously described [23]. The elastic modulus (storage modulus, G’) and viscous modulus (loss modulus, G′′) were directly extracted from the linear viscoelastic region of the amplitude sweep.

### Sputum microbiome analysis

DNA extraction and next generation sequencing of the v4 region of the 16S rRNA gene were performed as previously described [24].

### Sputum inflammation markers

IL-1β, IL-8 and tumor necrosis factor alpha (TNF-α) concentrations in cell-free sputum supernatants were quantified using cytometric bead array kits (BD Biosciences, San Diego, CA, USA) according to the manufacturer’s instructions. Free and membrane-bound NE activity were measured as previously described [25]. Protein intensities of other neutrophil serine proteases cathepsin G (CatG) and proteinase 3 (PR3), and of the antiproteases secretory leukocyte protease inhibitor (SLPI) and alpha-1-antitrypsin (AAT) were determined from sputum proteomics analysis.

### Sputum proteomics

Sputum samples were solubilized by adding 2% SDS buffer and incubated at 95°C for 10 minutes. Proteins were reduced, alkylated and treated with Benzonase. After single-pot solid-phase-enhanced sample-preparation protein cleanup proteins were treated with PNGase F followed by a tryptic digest. Cleaned-up peptide samples were measured on an EASY-nLC 1200 System coupled to an Orbitrap HF-X mass spectrometer (Thermo Fisher Scientific, Waltham, MA, USA) running on data-dependent acquisition mode as previously described [26].

### Statistical analysis

Data were analyzed using R 4.0.4. Clinical data are presented as mean and standard deviation and were tested by Student’s *t*-test. Data from measurements in sputum from healthy controls and patients with CF are presented as medians and interquartile range (IQR) and were compared by Mann-Whitney test. Changes between baseline and after initiation of ELX/TEZ/IVA were tested by Wilcoxon signed-rank test and are reported as median changes. Correction for multiple comparisons was performed using the Benjamini-Hochberg procedure. *P* < 0.05 was accepted to indicate statistical significance.

## Results

### Characteristics of study population

In total, 59 patients with CF were enrolled to assess anthropometry, spirometry, sweat chloride concentration, sputum viscoelastic properties, the sputum microbiome and inflammation markers, and the sputum proteome at baseline and 8 to 16 weeks after initiation of ELX/TEZ/IVA (supplementary figure S1). Four patients presented with pulmonary exacerbations at follow-up and were excluded from the analysis. Clinical characteristics of CF patients at baseline and after initiation of ELX/TEZ/IVA, and 10 age-matched healthy controls are summarized in Table 1. Patients with CF had a mean age of 30.1 ± 9.7 years, 54.5% were *F508del* homozygous and 45.5% had a previous dual CFTR modulator therapy with either LUM/IVA or TEZ/IVA (Table 1). Improvements in forced expiratory volume in one second (FEV_1_) %predicted, body mass index (BMI) and sweat chloride concentration (*P* < 0.001, Table 1) by ELX/TEZ/IVA therapy were consistent with previous studies [7-15, 27, 28]. *CFTR* genotypes are provided in supplementary table S1.

**Table 1.** Clinical characteristics of healthy controls and patients with cystic fibrosis at baseline and after initiation of elexacaftor/tezacaftor/ivacaftor.

| Clinical Characteristic | Healthy controls<br>mean (± SD) or n (%) | CF patients at<br>baseline<br>mean (± SD) or n (%) | CF patients with<br>ELX/TEZ/IVA<br>mean (± SD) or n (%) | Change between baseline<br>and ELX/TEZ/IVA<br>mean (± SD) | *P* value |
| --- | --- | --- | --- | --- | --- |
| Number of patients | 10 | 55 | 55 |  |  |
| Age (years) | 33.9 (± 4.8) | 30.1 (± 9.7) | 30.4 (± 9.7) | 0.3 (± 0.1) | |
| Pediatric patients (age < 18) | 0 (0%) | 6 (10.9%) |  |  |  |
| Sex (female) | 7 (70.0%) | 32 (58.2%) |  |  |  |
| Genotype |  |  |  |  |  |
| *F508del/F508del* | --- | 30 (54.5%) |  |  |  |
| *F508del/MF* | --- | 25 (45.5%) |  |  |  |
| No CFTR modulator | --- | 30 (54.4%) |  |  |  |
| TEZ/IVA therapy | --- | 18 (32.7%) |  |  |  |
| LUM/IVA therapy | --- | 7 (12.7%) |  |  |  |
| Pancreatic insufficiency (%) | --- | 55 (100.0%) |  |  |  |
| Sweat chloride (mmol/L) | --- | 91.0 (± 19.9) | 44.6 (± 23.0) | -45.7 (± 21.1) | <0.001 |
| FEV<sub>1</sub> %predicted | --- | 44.5 (± 23.2) | 53.2 (± 24.8) | 8.6 (± 7.7) | <0.001 |
| BMI (kg/m<sup>2</sup>) | 24.2 (± 3.2) | 19.8 (± 3.0) | 20.9 (± 2.8) | 1.1 (± 1.1) | <0.001 |

Definition of abbreviations: BMI = body mass index; ELX = elexacaftor; FEV<sub>1</sub> = forced expiratory volume in one second; IVA = ivacaftor;
LUM = lumacaftor; MF = minimal function mutation; SD = standard deviation; TEZ = tezacaftor. *P* value describes the change between baseline and elexacaftor/tezacaftor/ivacaftor therapy.

### ELX/TEZ/IVA improves viscoelastic properties of CF sputum

To study the effects of ELX/TEZ/IVA on sputum viscoelastic properties, we performed rheological measurements and determined the elastic modulus (G’), viscous modulus (G’’), and the mesh-pore size (ξ) of sputum samples from 10 healthy controls and 34 patients with CF at baseline and after initiation of therapy. In both groups, G’ predominated over G’’ indicating a gel-like behavior of healthy and CF sputum (figures 1a and 1b). In sputum from CF patients collected at baseline, G’ and G’’ were increased (*P* < 0.001) and the mesh-pore size was decreased (*P* < 0.001) compared to healthy controls (figure 1). Initiation of ELX/TEZ/IVA significantly reduced G’ by -6.3 Pa (IQR, -17.9 to 1.2; *P* < 0.01) and G’’ by -1.6 Pa (IQR, -3.6 to 0.5; *P* < 0.05), and increased mesh-pore size by 11.8 nm (IQR, -6.7 to 37.5; *P* < 0.05) in sputum from CF patients (figure 1). Compared to healthy controls, G’ and G’’ remained significantly elevated (*P* < 0.01) and mesh-pore size was still decreased (*P* < 0.01) in sputum from CF patients under ELX/TEZ/IVA therapy (figure 1).

**Figure 1.**
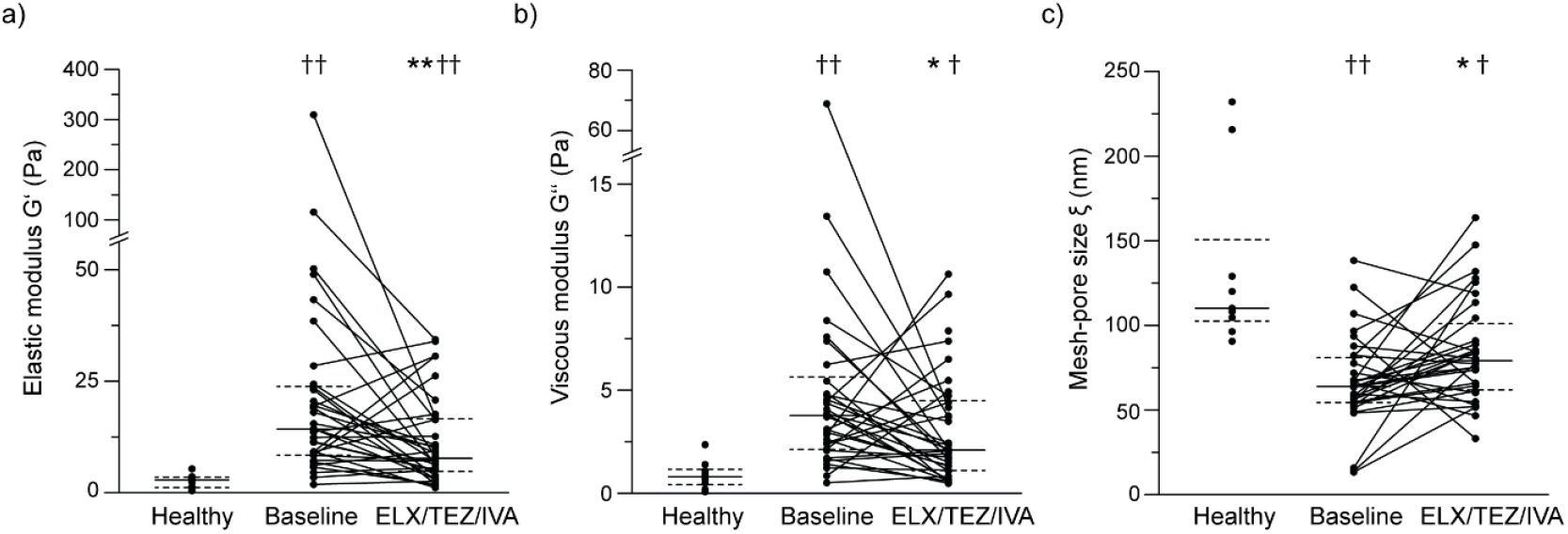
Elexacaftor/tezacaftor/ivacaftor (ELX/TEZ/IVA) therapy improves sputum viscoelastic properties in patients with cystic fibrosis (CF). (a-c) Measurements of the elastic modulus G’ (a), the viscous modulus G’’ (b), and the mesh-pore size ξ (c) of sputum from healthy controls (n = 10) and CF patients (n = 34) at baseline and after initiation of ELX/TEZ/IVA therapy. Solid lines represent the group median, dashed lines represent the 25th and 75th percentile. \**P* < 0.05 and \*\**P* < 0.01 compared with baseline; ^†^*P* ˂ 0.01 and ^††^*P* ˂ 0.001 compared with healthy.

### ELX/TEZ/IVA improves airway microbiome composition

To determine effects of ELX/TEZ/IVA on the airway microbiome, we performed 16S rRNA gene sequencing in 51 patients with CF at baseline and after initiation of therapy. At baseline, the most abundant genus were *Pseudomonas, Streptococcus, Prevotella* and *Veillonella* (figure 2a). *Pseudomonas* genus was detected in 67% of patients at baseline and in 61% of patients after initiation of ELX/TEZ/IVA (figure 2a). The total bacterial load did not change after initiation of ELX/TEZ/IVA (*P* = 0.567, figure 2b). However, ELX/TEZ/IVA improved the α-diversity (*P* < 0.001), evenness (*P* < 0.001), richness (*P* < 0.001) and dominance (*P* < 0.01) of the sputum microbiome of patients with CF (figures 2c-2f). Further, the relative abundance of *Pseudomonas aeruginosa* decreased from 10.8% (IQR, 0.0 to 48.7) at baseline to 2.6% (IQR, 0.0 to 43.6; *P* < 0.05) under ELX/TEZ/IVA (figure 2g). The cluster with high abundance of *Pseudomonas aeruginosa*, determined by hierarchical clustering based on the Morisita-horn distance between samples (supplementary figure S2), was reduced from 25% at baseline to 8% under ELX/TEZ/IVA, mainly towards the cluster with low abundance of *Pseudomonas aeruginosa*, but also towards clusters with commensal aerobes and anaerobes (figure 2h).

**Figure 2.**
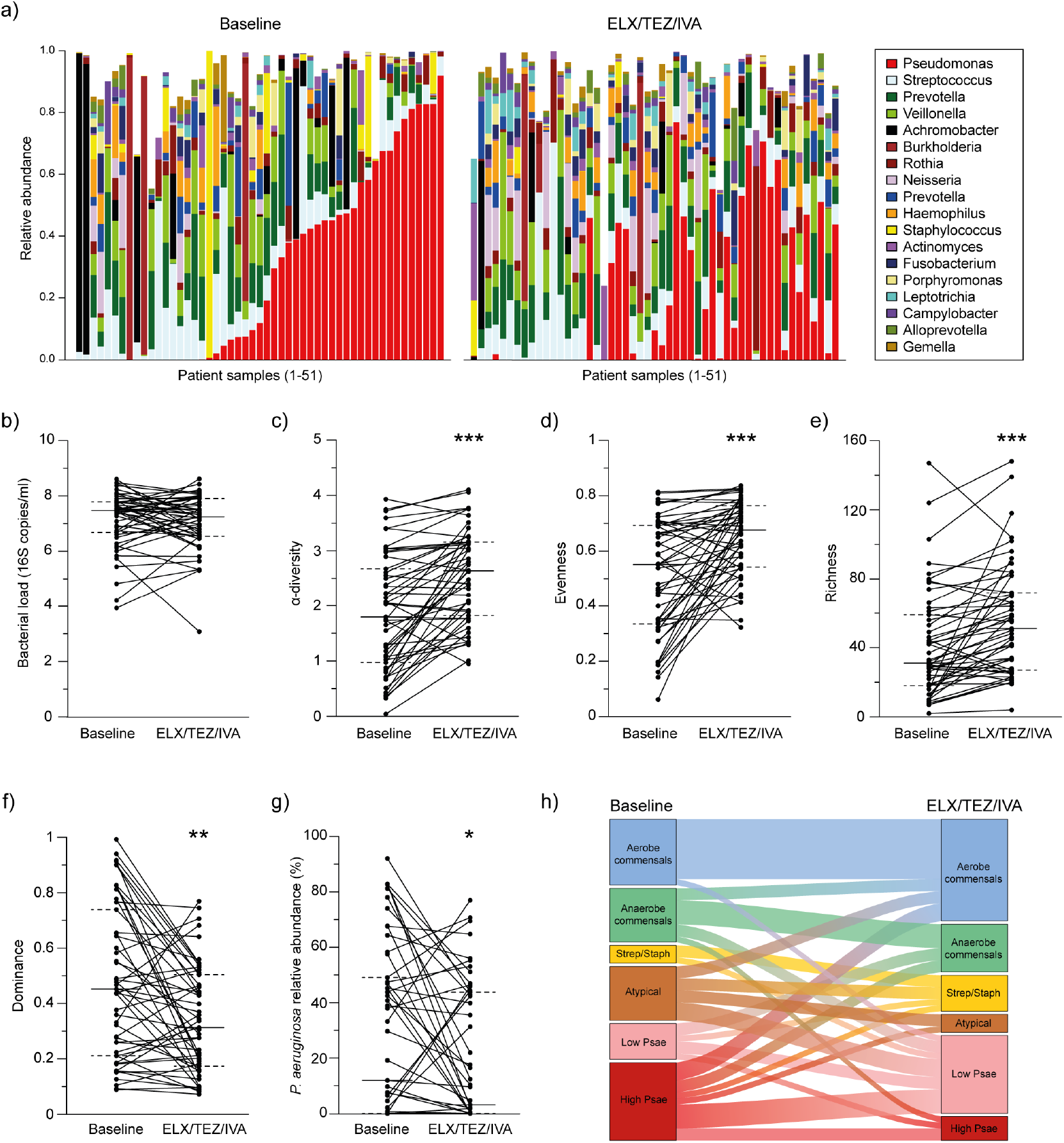
Elexacaftor/tezacaftor/ivacaftor (ELX/TEZ/IVA) improves the airway microbiome composition in patients with cystic fibrosis (CF). (a-e) Paired analysis of sputum microbiome at baseline and after initiation of ELX/TEZ/IVA therapy (n = 51). (a) Relative abundance of the most abundant genera (minimal abundance > 5 % in at least one sample) for each individual patient at baseline and after initiation of ELX/TEZ/IVA therapy. Samples are organized by relative abundance of the genus *Pseudomonas* at baseline. (b) Total bacterial load was determined from the number of 16S copies measured by qPCR (log 10 transformed). (c) α-diversity was calculated by the Shannon index. (d) Evenness was determined by the Pielou’s evenness index. (e) Richness was estimated by the number of ribosomal sequence variants observed. (f) Dominance was determined from the relative abundance of the most abundant amplicon sequence variant. (g) Relative abundance of the species *Pseudomonas aeruginosa* (*P. aeruginosa*). (h) Alluvial graphic depicting the proportions of different clusters dominated by aerobe commensals, anaerobe commensals, *Streptococcus* and *Staphylococcus* (Strep/Staph), atypical pathogens (atypical), and low or high abundance of *Pseudomonas aeruginosa* (low and high Psae) (clustering based on the Morisita-horn distance between samples, see supplementary figure S2) of the airway microbiome from CF patients at baseline and after initiation of ELX/TEZ/IVA therapy. Solid lines represent the group median, dashed lines represent the 25th and 75th percentile. \**P* < 0.05 and, \*\**P* < 0.01 and \*\*\**P* < 0.001 compared with baseline.

### ELX/TEZ/IVA reduces airway inflammation

To determine the effects of ELX/TEZ/IVA on airway inflammation, we measured IL-1β, IL-8 and TNF-α concentrations, i.e. key inflammation markers previously shown to be associated with CF lung disease progression [4], in sputum from 10 healthy controls and 41 CF patients at baseline and after initiation of ELX/TEZ/IVA. At baseline, IL-1β, IL-8 and TNF-α levels were elevated in CF compared to healthy sputum (*P* < 0.001; figure 3). ELX/TEZ/IVA significantly reduced IL-1β (−0.6 ng/ml, IQR, -2.3 to -0.1; *P* < 0.001) and IL-8 (−11.7 ng/ml, IQR, -36.5 to 11.2; *P* < 0.05) and led to a trend towards reduction of TNF-α (*P* = 0.092) (figure 3). Compared to healthy controls, IL-1β, IL-8 and TNF-α were still elevated in sputum from CF patients treated with ELX/TEZ/IVA (all *P* < 0.001; figure 3).

**Figure 3.**
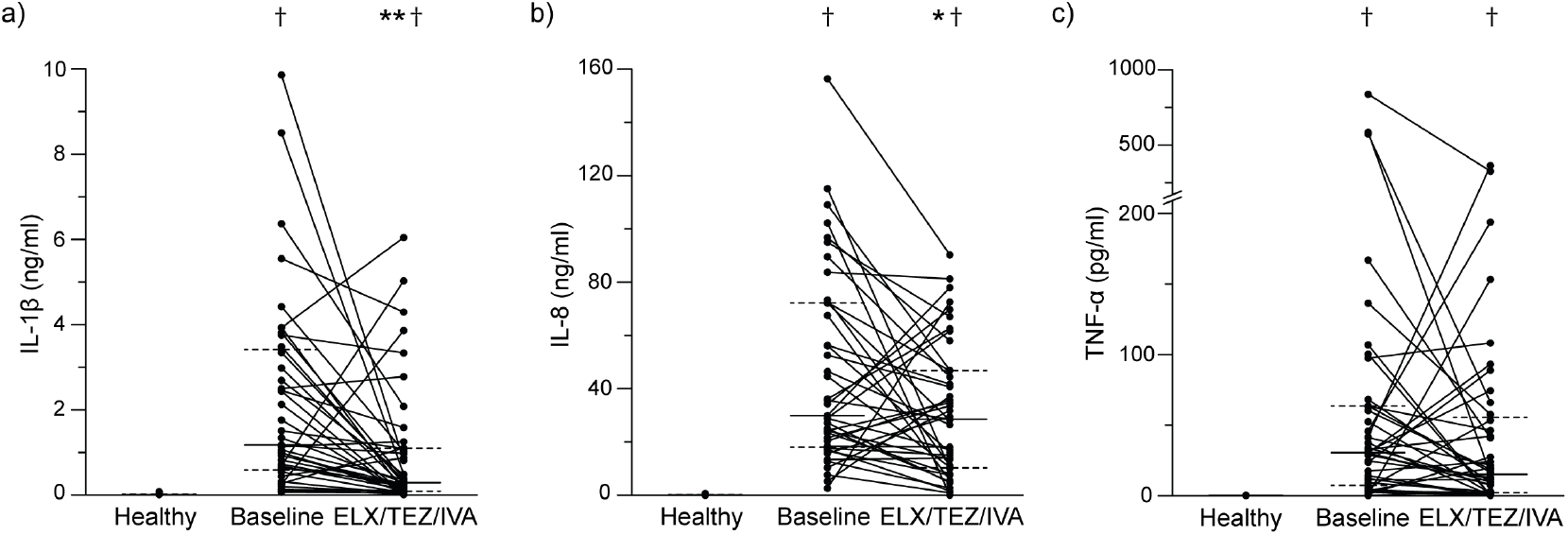
Elexacaftor/tezacaftor/ivacaftor (ELX/TEZ/IVA) therapy reduces airway inflammation in patients with cystic fibrosis (CF). (a-c) Measurements of interleukin (IL)-1β (a), IL-8 (b) and tumor necrosis factor alpha (TNF-α) in sputum from healthy controls (n = 10) and CF patients (n = 41) at baseline and after initiation of ELX/TEZ/IVA therapy. Solid lines represent the group median, dashed lines represent the 25th and 75th percentile. \**P* < 0.05 and \*\**P* < 0.001 compared with baseline; ^†^*P* ˂ 0.001 compared with healthy.

### ELX/TEZ/IVA reduces airway protease burden

Protease/antiprotease imbalance with increased NE activity in the airways plays an important role in the onset and progression of structural lung damage in CF [5, 6]. To determine the effects of ELX/TEZ/IVA on this protease/antiprotease imbalance in CF airways, we measured free NE activity in sputum and membrane-bound NE activity on sputum neutrophils, and determined the abundance of the neutrophil serine proteases CatG and PR3, as well as the antiproteases SLPI and AAT by label-free shotgun proteomics in sputum of healthy controls and CF patients at baseline and after initiation of therapy. At baseline, free and membrane-bound NE activity was elevated in CF compared to healthy sputum (*P* < 0.001; figures 4a and 4b). ELX/TEZ/IVA significantly reduced the activity of free NE (*P* < 0.001) and membrane-bound NE (*P* < 0.05) in sputum from CF patients (figures 4a and 4b). CatG and PR3 were also elevated in sputum of CF patients at baseline and were reduced after initiation of ELX/TEZ/IVA (*P* < 0.001; figures 4c and 4d). Measurements of antiproteases showed that SLPI was reduced (*P* < 0.05) and AAT was increased (*P* < 0.001) in sputum of CF patients at baseline compared to healthy controls (figures 4e and 4f). Under ELX/TEZ/IVA therapy SLPI showed a trend towards increase (*P* = 0.086) and AAT was significantly decreased in sputum from CF patients (*P* < 0.001; figures 4e and 4f). Compared to healthy controls, free and membrane-bound NE activity (*P* < 0.05), as well as CatG, PR3 and AAT levels (all *P* < 0.001) were still elevated in sputum from CF patients treated with ELX/TEZ/IVA (figure 4).

**Figure 4.**
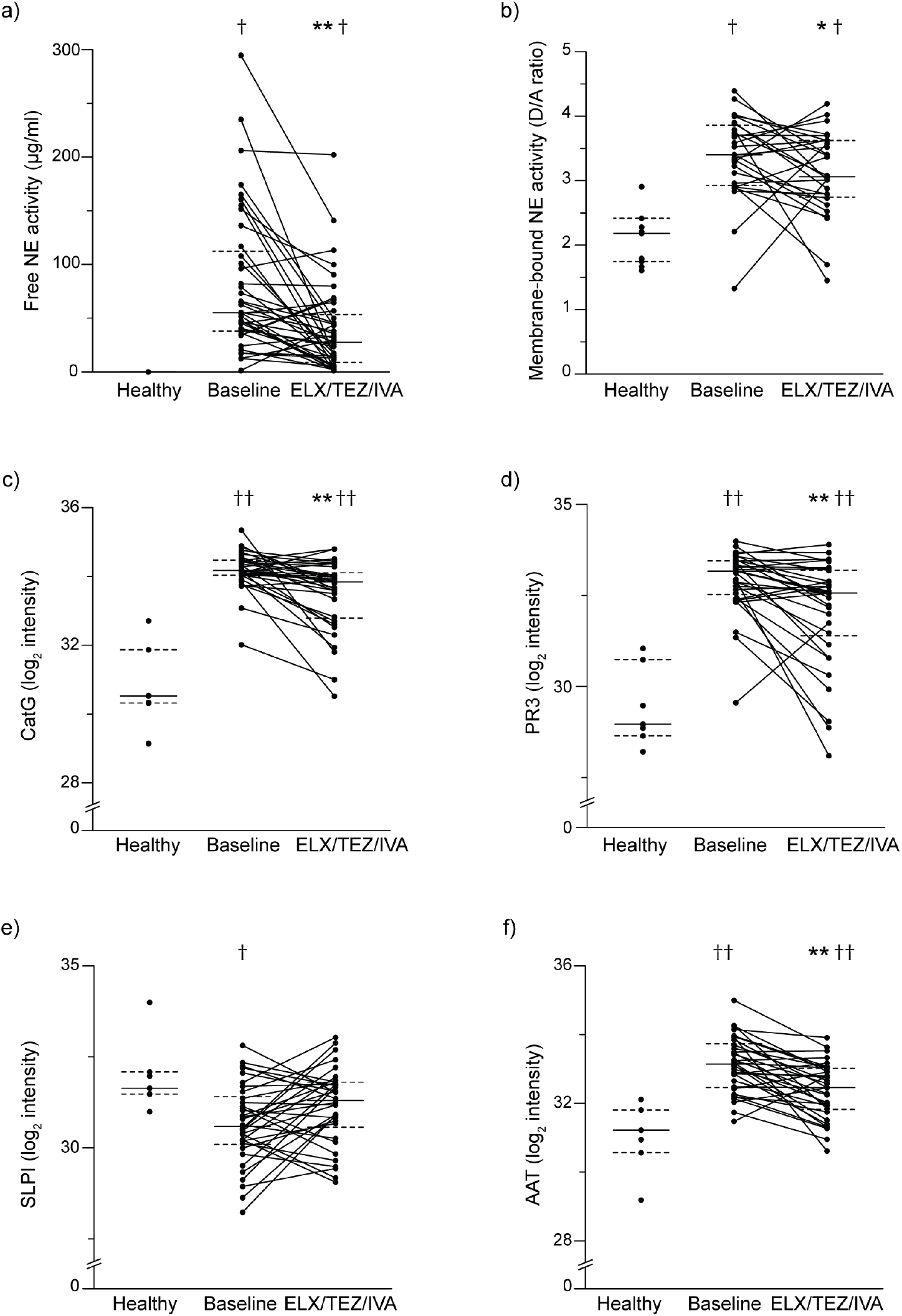
Elexacaftor/tezacaftor/ivacaftor (ELX/TEZ/IVA) therapy reduces airway protease burden in patients with cystic fibrosis (CF). (a-b) Measurements of free neutrophil elastase (NE) activity (a) and membrane-bound NE activity on neutrophils (b) in sputum from healthy controls (n = 10) and CF patients at baseline and after initiation of ELX/TEZ/IVA therapy (n = 41 for A and n = 28 for B). (c-f) Peptide intensity of the neutrophil serine proteases cathepsin G (CatG) (c) and proteinase 3 (PR3) (d), and the antiproteases secretory leukocyte peptidase inhibitor (SLPI) (e) and alpha-1-antitrypsin (AAT) (f) in sputum from healthy controls (n = 7) and CF patients (n = 34) at baseline and after initiation of ELX/TEZ/IVA therapy. Solid lines represent the group median, dashed lines represent the 25th and 75th percentile. \**P* < 0.05 and \*\**P* < 0.001 compared with baseline; ^†^*P* ˂ 0.05 and ^††^*P* ˂ 0.001 compared with healthy.

### ELX/TEZ/IVA shifts the CF sputum proteome towards healthy

Finally, we performed label-free shotgun proteomics of sputum from 7 healthy controls and 34 patients with CF at baseline and after initiation of ELX/TEZ/IVA as an unbiased approach to assess the effects of ELX/TEZ/IVA on abnormalities of the CF sputum proteome. In total, 2742 proteins were quantified, of which 1474 (54%) were significantly different (cutoff *P* < 0.01) between sputum from healthy controls and patients with CF at baseline. After initiation of ELX/TEZ/IVA, 472 proteins (17%) were differentially expressed compared to baseline. 1046 (38%) of all detected proteins were significantly different between all three groups and were further grouped into two major clusters using hierarchical clustering (figure 5a). Cluster 1 contains proteins that are expressed at lower levels in CF compared to healthy sputum. Gene set enrichment analysis of gene ontology (GO) terms showed that these proteins are involved in processes such as protein targeting to the endoplasmic reticulum (ER) and the plasma membrane (figure 5a). Cluster 2 includes proteins that are expressed at higher levels in sputum from CF patients compared to healthy individuals, and is enriched in proteins linked to immunological and inflammatory processes, such as neutrophil activation, phagocytosis and apoptosis (figure 5a). After initiation of ELX/TEZ/IVA, both proteome clusters in CF sputum shifted towards the expression pattern of healthy controls (figure 5a). Gene set enrichment analysis of GO terms showed that ELX/TEZ/IVA treatment partially restored the amount of proteins that showed a lower abundance in sputum of CF patients at baseline, and were assigned to signal recognition particle (SRP)-dependent co-translational protein targeting to the membrane, protein targeting to the ER and epithelial cell differentiation (figure 5b). Furthermore, proteins involved in inflammatory processes, such as granulocyte activation, neutrophil mediated immunity including neutrophil activation and degranulation, and phagocytosis, showed increased levels in sputum of patients with CF compared to healthy controls and were reduced after initiation of ELX/TEZ/IVA (figure 5b). The complete list of significantly different GO terms is provided in the online supplement (supplementary figure S3).

**Figure 5.**
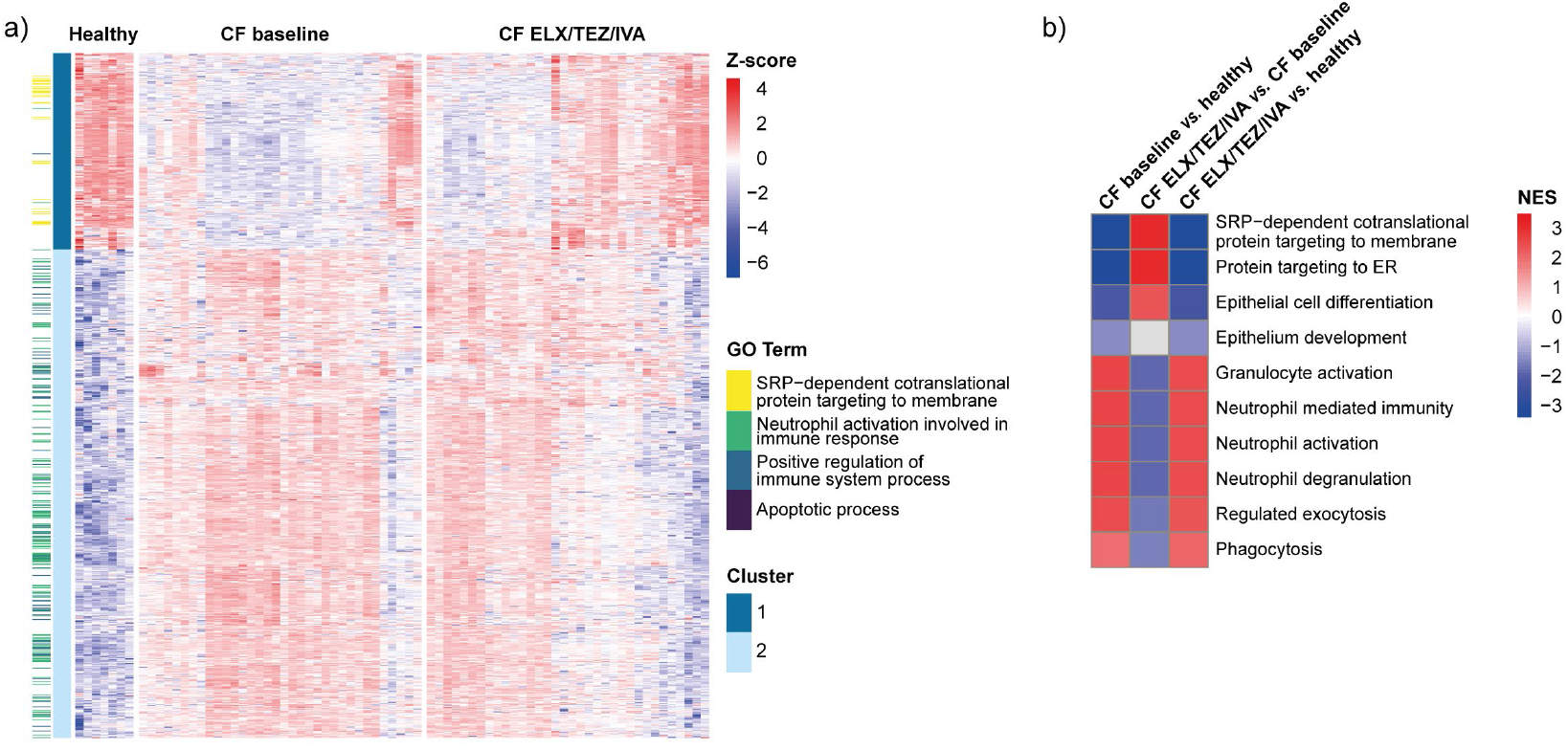
Elexacaftor/tezacaftor/ivacaftor (ELX/TEZ/IVA) therapy shifts the sputum proteome of patients with cystic fibrosis (CF) towards healthy. (a) Heatmap of significantly differentially expressed proteins (cutoff *P* < 0.01) in sputum from healthy controls (n = 7) and CF patients (n = 34) at baseline and after initiation of ELX/TEZ/IVA therapy clustered by hierarchical clustering in a cluster with downregulated proteins (cluster 1) and a cluster with upregulated proteins (cluster 2) in CF patients at baseline compared to healthy controls. Proteins that contribute most to representative enriched gene ontology (GO) terms are highlighted in the first column. (b) Gene set enrichment analysis of GO terms in CF sputum at baseline compared to healthy controls (first column), in CF sputum after initiation of ELX/TEZ/IVA compared to baseline (second column) and in CF sputum after initiation of ELX/TEZ/IVA compared to healthy controls (third column) ordered by normalized enrichment score (NES). SRP = signal-recognition particle; ER = endoplasmatic reticulum.

## Discussion

This is the first study that assessed the effects of the triple combination CFTR modulator therapy with ELX/TEZ/IVA on sputum viscoelastic properties, the airway microbiome and chronic airway inflammation in adolescent and adult patients with CF with at least one *F508del* allele. In this post-approval study in a real-world setting, the clinical responses to ELX/TEZ/IVA on lung function, BMI and sweat chloride concentration were in line with results of controlled clinical trials and other observational studies (Table 1) [7-15, 27, 28]. Our rheological measurements show that treatment with ELX/TEZ/IVA improved the viscoelastic properties by reducing the elastic (G’) and viscous (G’’) moduli of sputum from patients with CF (figure 1). Further, initiation of ELX/TEZ/IVA led to an improvement in the composition of the airway microbiome, including an increase in α-diversity and a reduction in the relative abundance of *Pseudomonas aeruginosa*, and to a reduction in airway inflammation and protease burden in patients with CF (figures 2 and 4). In addition, using proteome analysis as an unbiased approach, we found that alterations in the sputum proteome of CF patients were shifted towards the sputum proteome of healthy controls under ELX/TEZ/IVA therapy (figure 5). However, by comparison to sputum from healthy controls, our data also demonstrate that sputum rheology, inflammation markers and protease burden, as well as proteome signatures remain abnormal in CF patients under ELX/TEZ/IVA therapy. Collectively, our results provide novel insights into the impact of improvement of CFTR function by ELX/TEZ/IVA on mucus properties, chronic airway infection and inflammation, and protease burden that constitute key risk factors in the onset and progression of CF lung disease.

Abnormal viscoelasticity of airway mucus, caused by increased mucin concentration due to airway surface dehydration, increased concentration of DNA released from inflammatory cells, and increased disulfide crosslinking of mucins caused by reactive oxygen species generated in neutrophilic airway inflammation, is a key feature of CF that correlates with the severity of lung function impairment as well as airway infection and inflammation in patients [1-3, 29-32]. Previous studies showed that LUM/IVA improves viscoelastic properties of CF mucus [33], and that ELX/TEZ/IVA reduces mucin concentration and improves mucociliary transport in CF human bronchial epithelial cells *in vitro* [34]. We show for the first time that ELX/TEZ/IVA improves viscoelastic properties of sputum from patients with CF *in vivo* (figure 1). Despite substantial improvement in the elastic (G’) and viscous (G’’) moduli, they remained elevated compared to sputum from healthy controls indicating that the level of rescue of CFTR function achieved by ELX/TEZ/IVA may not be sufficient for full restoration of mucus properties in patients with CF.

Pathological CF mucus causing impaired mucociliary clearance sets the stage for chronic polymicrobial infection and inflammation, and improvement of viscoelastic properties of the mucus has therefore the potential to limit these abnormalities in CF airways [1, 2, 4, 5, 35]. However, studies investigating the effects of prior CFTR modulator drugs on airway infection and inflammation reported mixed results [36-41]. One study investigating the effects of LUM/IVA that is less effective in restoring CFTR function than ELX/TEZ/IVA showed a reduction in sputum total bacterial load and an improvement in α-diversity of the microbiome accompanied by improved IL-1β, while another study observed no change in α-diversity, but a trend towards a reduction in the relative abundance of *Pseudomonas aeruginosa* [19, 36, 37]. Treatment with IVA in patients with a *G551D* mutation, which is similarly effective in restoring CFTR function as ELX/TEZ/IVA in patients with at least one *F508del* allele [15, 17], led to a reduction in the relative abundance of *Pseudomonas aeruginosa* and an increase in the α-diversity of the airway microbiome, which was also associated with improvements in airway inflammation [38-41]. In our study, we found a similar improvement of the microbiome composition including a reduction in the relative abundance of *Pseudomonas aeruginosa* in patients with one or two *F508del* alleles under ELX/TEZ/IVA therapy (figure 2). These findings are consistent with a small study showing an increase in the microbiome α-diversity and evenness, and a trend towards the reduction of *Pseudomonas sp*. with ELX/TEZ/IVA [21]. Of note, long-term studies with IVA also reported a rebound of improvements of the microbiome after the first year of treatment [39]. Therefore, such long-term studies will be necessary to investigate the microbiome trajectory and determine if the beneficial effects of ELX/TEZ/IVA observed in our study persist over time.

In addition to these improvements of the sputum microbiome, initiation of ELX/TEZ/IVA also led to improvements in key inflammation markers such as IL-1β and IL-8 that have been implicated in CF lung disease progression [4] (figure 3). The effects on these proinflammatory cytokines observed in our study are in line with those reported for IVA in CF patients with a *G551D* mutation, and greater than those observed for LUM/IVA in *F508del* homozygous patients [37-40]. Besides elevated proinflammatory cytokines, neutrophilic airway inflammation is associated with sustained proteolytic activity that is caused by a protease/antiprotease imbalance and constitutes a key driver of lung damage and disease progression in CF [4-6]. In this context, increased activity of NE, but also of the other neutrophil serine proteases CatG and PR3, are important contributors to the protease burden of the CF lung [4-6, 42-46]. In our study, we observed a decrease in NE activity and reduced sputum levels of CatG and PR3 after initiation of ELX/TEZ/IVA (figure 4). These effects on neutrophil serine proteases were associated with improvements of AAT and SLPI levels [4, 6, 42, 47], suggesting an overall improvement of protease/antiprotease imbalance in the lungs of CF patients under ELX/TEZ/IVA therapy. However, long-term studies will be required to determine the impact of this reduction in protease burden on progression of structural lung damage in patients with CF.

Our studies of established markers of CF airway inflammation were complemented by sputum proteomics analyses as an unbiased approach to assess effects of ELX/TEZ/IVA on molecular pathways altered in CF lung disease. In line with reduced inflammation markers and neutrophil serine proteases, we observed a global reduction of proteins linked to inflammatory processes, such as neutrophil activation/degranulation and phagocytosis after initiation of ELX/TEZ/IVA (figure 5). Further, proteins involved in airway epithelial cell differentiation that were reduced in sputum from CF patients at baseline were upregulated under ELX/TEZ/IVA, which may reflect improved epithelial homeostasis. In addition, we found that ELX/TEZ/IVA increased the abundance of proteins related to SRP-dependent co-translational protein targeting to the ER that were previously identified as high-confidence members of the F508del-CFTR interactome (figure 5 and supplementary figure S3) [48]. We speculate that pharmacologically corrected F508del-CFTR may have specific binding partners during processing and trafficking that are only part of the CFTR-interactome when the protein is folded and not prematurely degraded. This notion is supported by a recent study showing large differences in the wild-type and F508del-CFTR interactomes [49].

This real-world observational study has limitations. First, we included patients with a large range in age and baseline lung function, possibly influencing the observed treatment effects. Second, we focused on short-term treatment effects and all measurements were performed in a strictly paired fashion in CF patients that expectorated sputum at baseline and 8 to 16 weeks after initiation of ELX/TEZ/IVA, biasing our study towards patients with more advanced lung disease. In this patient population, despite the substantial improvements of sputum outcomes observed under ELX/TEZ/IVA, sputum viscoelasticity, inflammation markers and protease burden remained elevated, and abnormal proteome signatures were only partially restored compared to healthy controls. Whether long-term treatment in patients with established lung disease, or early initiation of ELX/TEZ/IVA therapy in children with CF with preserved lung function [50], can achieve a more complete resolution of abnormal mucus properties, airway dysbiosis and inflammation remains to be determined in future studies. However, our data support the use of the sputum outcome measures utilized in our study for the assessment of effects of novel therapeutic interventions targeting increased mucus viscoelasticity, airway infection and inflammation in patients with CF and possibly other muco-obstructive lung diseases.

In summary, our study demonstrates that ELX/TEZ/IVA therapy leads to improvements in sputum viscoelastic properties, chronic airway infection and inflammation as well as abnormalities in airway proteome signatures in patients with CF 12 years and older with one or two *F508del* alleles. Our data on residual disease activity under this highly effective CFTR modulator therapy also indicate that additional therapeutic strategies may be needed to control airway infection and inflammation in adolescent and adult CF patients with chronic lung disease.

## Supporting information

Oline Supplement

## Data Availability

Pseudonomyzed microbiome data and scripts are available upon reasonable request to the authors. Due to data safety constrictions, for proteomics only pooled data will be available.

## Acknowledgements

The authors thank the patients with CF for their participation in this study; M. Daniltchenko, M. Drescher, S. Mayer, A. Rohrbach, K. Seidel and J. Tattersall-Wong for excellent technical assistance; C. Labitzke for excellent documentation; and our clinical colleagues for clinical care of study participants.

