## Supplementary material for "Effects of Elexacaftor/Tezacaftor/Ivacaftor on Sputum Viscoelastic Properties, Airway Infection and Inflammation in Patients with Cystic Fibrosis": Oline Supplement

**Online Supplementary Material**

### Supplementary methods

#### Study design and participants

Patients were eligible to participate if they were at least 12 years old, compound-heterozygous for *F508del* and a minimal function mutation or homozygous for *F508del*, had no prior exposure to ELX/TEZ/IVA and were willing to remain on a stable medication regimen including ELX/TEZ/IVA according to the patient labeling and the prescribing information for the duration of study participation. Exclusion criteria were an acute respiratory infection or pulmonary exacerbation at baseline. Sputum rheology, microbiome analysis, cytokine measurements and proteomics were assessed if patients provided sputum at baseline and 8 to 16 weeks after initiation of therapy with the approved dose of ELX 200 mg and TEZ 100 mg every 24 hours in combination with IVA 150 mg every 12 hours (supplementary Figure S1). Due to limited amount of sputum not all measurements could be performed with every sputum sample.

#### Lung function

Spirometry was performed and forced expiratory volume in one second (FEV1), was determined according to ATS/ERS standards [1]. Percent predicted results were based on equations of the global lung initiative [2].

#### Sweat chloride concentration (SCC)

Sweat tests were performed according to the German national diagnostic guideline [3] and the guidelines of the Clinical and Laboratory Standards Institute [4]. Sweating was stimulated by pilocarpine iontophoresis and samples were collected with the Macroduct® system (Model 3700, Wescor, Logan UT, USA). Sweat chloride concentration was measured in a minimum volume of 30 µl using a chloridometer (KWM 20 Chloridometer, Kreienbaum, Langenfeld, Germany).

**Sputum rheology**

Native sputum samples were immediately put on ice after expectoration and sputum was separated from saliva and possible debris by gentle aspiration with a pipette. Rheological measurements were immediately performed with a cone-and plate rheometer (Kinexus Pro+, Netzsch GmbH, Selb, Germany). All experiments were performed using a stainless-steel cone-plate geometry (cone-diameter 20 mm, cone-angle 1°). Samples were transferred onto the lower static plate of the rheometer with a non-electrostatic spatula. After loading and between the performed sequences, the samples were equilibrated for five minutes at 25°C to ensure full temperature equilibration and sufficient network relaxation. Each measurement included an amplitude sweep and a frequency sweep downwards. The amplitude sweep was performed at a fixed frequency of 1 Hz and covered a range of shear deformation γ between 0.01 - 100%. The frequency sweep was conducted at a fixed shear deformation of 2% and covered a frequency range from 10 to 0.1 Hz. The elastic modulus (storage modulus, G') and viscous modulus (loss modulus, G″) were directly extracted from the linear viscoelastic region of the amplitude sweep. To estimate an effective mesh-pore size ξ of the mucin network, we used the following formula as previously described [5]:

$$\xi = \left(\frac{\boldsymbol{k_B} \cdot \boldsymbol{T}}{\boldsymbol{G}}\right)^{\frac{1}{3}}$$

with kB being the Boltzmann constant, T the absolute temperature and G the shear modulus, which we approximated from the G' value of the frequency sweep at 1 Hz.

**Sputum microbiome analysis**

DNA extractions were performed using the QIAamp Mini Kit (Qiagen, Hilden, Germany). Protease solution (7.2 mAU) and 200 µl of Buffer AL were added to 200 µl of the sputum sample followed by a 15 sec vortex. Samples were incubated at 56°C for 10 min and then purified according to the manufacturer's protocol. DNA was eluted by adding 100 µl of buffer AE to the column, incubation for 1 min at room temperature and centrifugation at 6000 x g for 1 min. Negative controls were performed by doing the extraction without clinical samples. DNA was amplified using universal bacterial primers flanking the V4 region (515F and 806R from) [6]. Each primer was tagged with an individual barcode (each barcode had at least 3 nucleotides differences to the others) to assign the sequences to the samples. PCR reactions were performed in 25 µl volumes composed of Q5 High-Fidelity 1x Master Mix (New England Biolabs GmbH, Frankfurt am Main, Germany), 25 pmol of each primer and 2 µl of DNA. The thermal cycler (Primus 25, Peqlab Biotechnologie GmbH, Erlangen, Germany or FlexCycler², Analytik Jena AG, Jena, Germany) conditions were: a first denaturation at 94°C for 3 min, 30 amplification cycles (94°C for 45 sec, 50°C for 1 min and 72°C for 1.5 min) and a final extension at 72°C for 10 min. Negative controls were performed using the negative control from the extraction step and sterile water as template. For each run of sequencing (pool of 95 samples), an internal control was performed by amplifying a mock community sample containing genomic DNA from 20 bacterial strains in equimolar (even) ribosomal RNA operon counts (HMD-782D, BEI resources, ATCC, Manassas, VA, USA). PCR products were checked on Qiaxcel device for the presence of amplicons. Amplicons were then ligated to Illumina by PCR and purified using Agencourt AMPure XP beads (Beckman Coulter, Krefeld, Germany) following the manufacturer's instructions. Purified products were checked for concentration using Quant-iT™ PicoGreen™ dsDNA Assay Kit (Thermo Fisher Scientific, Waltham, MA, USA) following manufacturer's instructions. An equimolar mix of all PCR products was pooled and paired-end sequenced on an Illumina Miseq sequencing system (Illumina, San Diego, CA, USA) with 600 cycles. Both negative controls were negative on the gel and library preparation resulted in no usable reads (percentage of reads in the run < 0.005% and 0 reads remained after quality control and chimera removal).

Raw reads were processed using the Dada2 package (v.1.12.1) to produce ribosomal sequence variants (RSVs) [7]. The reads were filtered and trimmed using the following parameters: no ambiguities allowed; one error per read allowed; truncation of reads at the first position with a quality score of less than two. Chimeras were removed following the default parameters. Taxonomic assignment of RSVs was performed using the 'assignTaxonomy' function in Dada2, which utilized the Ribosomal Database Project Naïve Bayesian Classifier method and training sets constructed from the Silva v138 database [8, 9]. To detect differences in the abundance of RSVs, the DESeq2 algorithm was used [10]. Sequencing data and script are available in figshare (https://figshare.com/s/74fdc6850bfe9e734ec9, 10.6084/m9.figshare.20435697). For each sample, the bacterial communities were characterized by calculating α-diversity (Shannon index), species evenness (Pielou index), richness (number of RSV observed), and dominance (relative abundance of the most abundant RSV). Pulmotypes were determined by using hierarchical clustering on a tree based on the β-diversity using Ward method and using the rule of the first SE max to determine the optimal amount of cluster/pulmotype (aerobe and anaerobe commensals, atypical, *Streptococcus*/*Staphylococcus* (Strep/Staph), low and high *Pseudomonas aeruginosa* (low Psae and high Psae)) (supplementary figure S2).

The number of 16S copies was quantified by quantitative PCR (qPCR) using Unibac primer (forward: 5′-TGG AGC ATG TGG TTT AAT TCG A-3′; reverse: 5′-TGC GGG ACT TAA CCC AAC A-3′). PCR reactions were performed in 15 µl volumes composed of 1x SYBR Green Master Mix (Life technology, Darmstadt, Germany), 50 pmol of each primer and 2 µl of DNA (or plasmid DNA standards). The thermal cycler conditions were: a first denaturation at 95°C for 20 sec, 40 amplification cycles (95°C for 3 sec, 60°C for 30 sec) and two final steps at 95°C for 15 sec and 60°C for 1 minute followed by melt curve analysis for specificity control. All reactions were performed in duplicates in a StepOnePlus Real-time PCR system (Applied Biosystems, Foster City, CA, USA). Quantification of the 16S number of copies was performed by comparison to the cycle threshold value of a plasmid DNA standard, which had been quantified by spectrophotometry.

**Sputum inflammation markers**

Cells were isolated from sputum after treatment with 10% (v/v) sputolysin (Calbiochem, Darmstadt, Germany). The concentrations of interleukin (IL)-1 β, IL-8 and tumor necrosis factor alpha (TNF-α) in cell-free sputum supernatants were determined by cytometric bead array kits according to the manufacturer's instructions (BD Biosciences, San Diego, CA, USA). Membrane-bound neutrophil elastase (NE) activity on neutrophils was quantified by using flow cytometry in combination with the lipidated Förster resonance energy transfer (FRET) probe NEmo-2E (Sirius Fine Chemicals, Bremen, Germany) as previously described [11, 12]. Free NE activity was measured in cell-free sputum supernatants using the FRET reporter NEmo-1 (Sirius Fine Chemicals, Bremen, Germany). Kinetic assays were performed at 25°C using a fluorescence microplate reader (SpectraMax iD5, Molecular Devices, San Jose, CA, USA) and reporter cleavage was recorded over time ($\lambda_{Excitation}$ = 354 nm, $\lambda_{Emission\ Donor}$ = 400 nm and $\lambda_{Emission\ Acceptor}$ = 490 nm). Subsequently, NE activity was determined by calculating the ratio of donor to acceptor fluorescence (D/A ratio) and applying an enzyme standard curve as previously described [12].

**Sputum proteomics**

Sputum samples were inactivated and proteins solubilized by adding sodium dodecyl sulfate (SDS) buffer (4% SDS, 100 mM Tris-HCl pH 8, 1 mM EDTA, 150 mM NaCl) in a 1:1 volume to volume ratio, followed by an incubation at 95°C for 10 min. After measuring the protein concentration using a BCA assay, 100 µg protein were reduced and alkylated with 10 mM dithiothreitol (DTT) and 40 mM chloroacetamide at 95°C for 10 min. Subsequently, samples were treated with benzonase (25U, Merck, Darmstadt, Germany) for 15 min and protein clean-up was performed using the single-pot solid-phase-enhanced sample-preparation (SP3) protocol [13]. Protein containing beads were resuspended in 50 mM ABC buffer, treated with 2 µg peptide-N-glycosidase F (PNGase F) (NEB, Ipswich, MA, USA) for 1 hour at 37°C and subsequently digested with trypsin (Promega, Madison, WI, USA) and lysyl endopeptidase (LysC) (Fujifilm Wako Pure Chemical Corporation, Richmond, VA, USA) at a 1:50

enzyme:substrate ratio overnight at 37°C. The peptide containing supernatant was collected and desalted using C18 stage tips [14]. 2 µg of the peptides samples were measured using a 200 min gradient on an EASY-nLC 1200 System coupled to an Orbitrap HF-X mass spectrometer (Thermo Fisher Scientific, Waltham, MA, USA) running on data dependent acquisition (DDA) mode as previously described [15].

Raw data were analyzed using MaxQuant software package (Ver. 1.6.3.4; Max Planck Institute of Biochemistry, Martinsried, Germany) and a decoy human UniProt database (2020-06) [16]. Variable modifications of oxidation (M), N-terminal acetylation, deamidation (N, Q) and fixed modification of carbamidomethyl cysteine were selected. The false discovery rate (FDR) was set to 1% for peptide and protein identifications. Unique and razor peptides were considered for quantification. "Match between runs" and label-free quantitation (LFQ) algorithm were applied. MaxQuant protein groups data were filtered by removing reverse hits, proteins only identified by site and potential contaminants. Data were further filtered for proteins identified by at least two peptides or at least 5 MS/MS counts with an Andromeda score above 20. Outlier patient samples, defined by number of proteins identified (< than 1200) and principal component analysis, were excluded. Proteins identified in at least 50% of the remaining patient samples were considered for further analysis. Missing values were replaced by random values from a normal distribution with a width of 0.3 and a down shift of 1.8. Cathepsin G (CatG), proteinase 3 (PR3), secretory leukocyte protease inhibitor (SLPI) and alpha-1-antitrypsin (AAT) peptide intensities were calculated from log2-transformed non-imputed LFQ intensities. For statistical analysis two-sample or paired moderated t-testing and moderated F testing [17] were applied. *P*-values were adjusted using the Benjamini-Hochberg method and cutoffs of either 0.05 or 0.01 were chosen. For gene set enrichment analysis (GSEA) the clusterProfiler package was used [18]. Only gene ontology (GO) terms of biological processes with a minimum size of 50 and a maximum size of 500 were considered. Raw data and scripts are available on the Proteomics Identifications Database PRIDE.

### Supplementary tables

**Supplementary table S1.** List of cystic fibrosis transmembrane conductance regulator genotypes of patients with cystic fibrosis.

| First allele | Second allele | Number of patients |
|---|---|---|
| *F508del* | *F508del* | 30 |
| *F508del* | *G542X* | 3 |
| *F508del* | *N1303K* | 3 |
| *F508del* | *R347P* | 3 |
| *F508del* | *1078delT* | 2 |
| *F508del* | *R553X* | 2 |
| *F508del* | *1525-1G>A* | 1 |
| *F508del* | *1717-1G>A* | 1 |
| *F508del* | *2721del11* | 1 |
| *F508del* | *2790-2A>G* | 1 |
| *F508del* | *2991del32* | 1 |
| *F508del* | *CFTRdele17a,17b* | 1 |
| *F508del* | *CFTRdele2,3* | 1 |
| *F508del* | *M1101K* | 1 |
| *F508del* | *Q39X* | 1 |
| *F508del* | *R1158X* | 1 |
| *F508del* | *R709X* | 1 |
| *F508del* | *W1282X* | 1 |

**Supplementary figures**

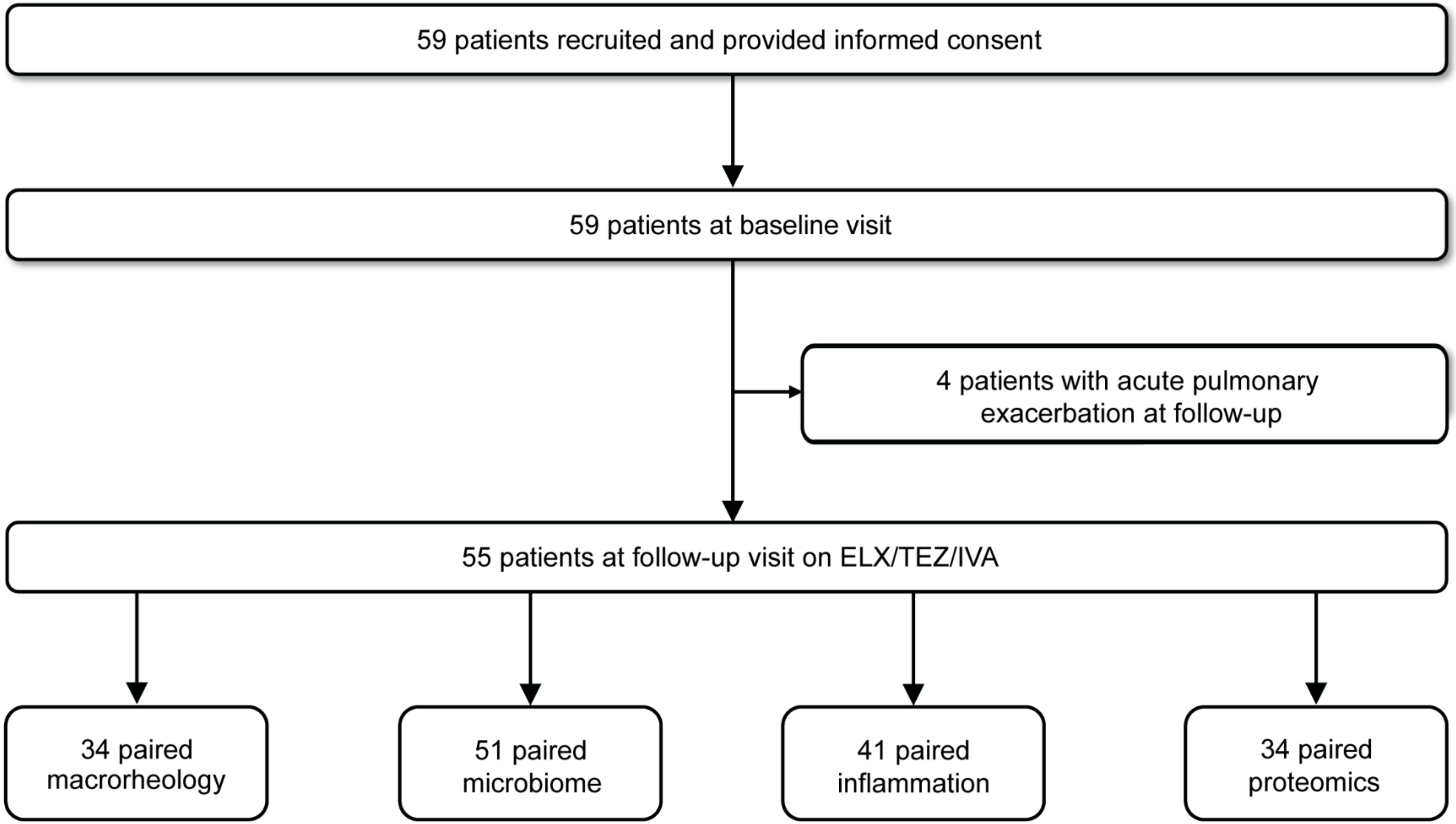


**Supplementary figure S1.** Flow chart of recruited patients with cystic fibrosis at baseline and at follow-up after initiation of elexacaftor/tezacaftor/ivacaftor (ELX/TEZ/IVA) therapy.

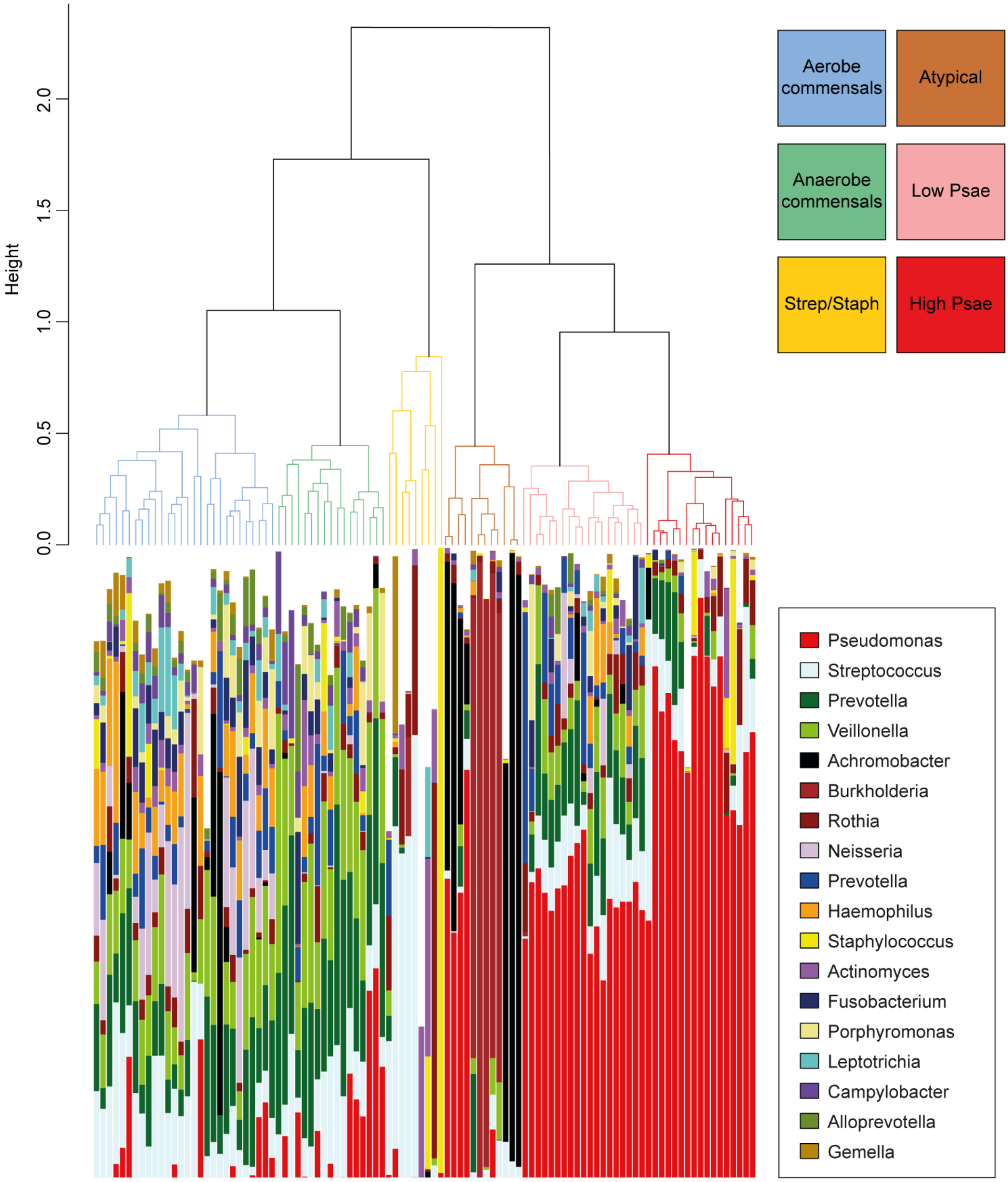


**Supplementary figure S2.** Hierarchical clustering dendrogram of sputum microbiota from patients with cystic fibrosis. Commensal species are dominant in two clusters with a switch between aerobic and anaerobic species dominance (aerobe and anaerobe commensals). The other clusters are characterized by the dominance of pathogenic species, such as *Streptococcus* and *Staphylococcus* (Strep/Staph), low or high abundance of *Pseudomonas aeruginosa* (low and high Psae) or more atypical pathogens like *Achromobacter*, *Burkholderia* or *Rothia* (atypical).

a)

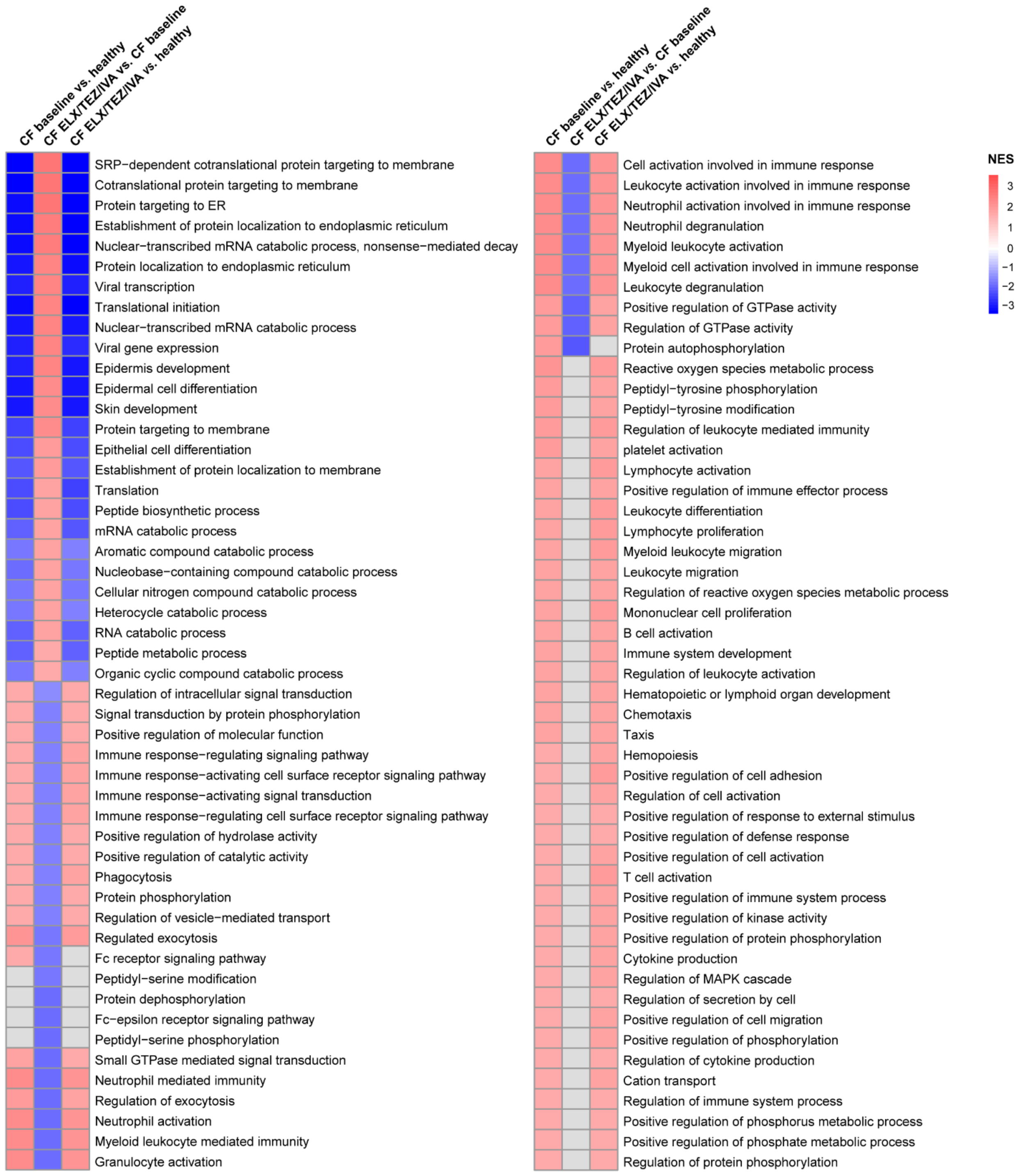
CF baseline vs. healthy
CF ELX/TEZ/IVA vs. CF baseline
CF ELX/TEZ/IVA vs. healthy
SRP−dependent cotranslational protein targeting to membrane
Cotranslational protein targeting to membrane
Protein targeting to ER
Establishment of protein localization to endoplasmic reticulum
Nuclear−transcribed mRNA catabolic process, nonsense−mediated decay
Protein localization to endoplasmic reticulum
Viral transcription
Translational initiation
Nuclear−transcribed mRNA catabolic process
Viral gene expression
Epidermis development
Epidermal cell differentiation
Skin development
Protein targeting to membrane
Epithelial cell differentiation
Establishment of protein localization to membrane
Translation
Peptide biosynthetic process
mRNA catabolic process
Aromatic compound catabolic process
Nucleobase−containing compound catabolic process
Cellular nitrogen compound catabolic process
Heterocycle catabolic process
RNA catabolic process
Peptide metabolic process
Organic cyclic compound catabolic process
Regulation of intracellular signal transduction
Signal transduction by protein phosphorylation
Positive regulation of molecular function
Immune response−regulating signaling pathway
Immune response−activating cell surface receptor signaling pathway
Immune response−activating signal transduction
Immune response−regulating cell surface receptor signaling pathway
Positive regulation of hydrolase activity
Positive regulation of catalytic activity
Phagocytosis
Protein phosphorylation
Regulation of vesicle−mediated transport
Regulated exocytosis
Fc receptor signaling pathway
Peptidyl−serine modification
Protein dephosphorylation
Fc−epsilon receptor signaling pathway
Peptidyl−serine phosphorylation
Small GTPase mediated signal transduction
Neutrophil mediated immunity
Regulation of exocytosis
Neutrophil activation
Myeloid leukocyte mediated immunity
Granulocyte activation
CF baseline vs. healthy
CF ELX/TEZ/IVA vs. CF baseline
CF ELX/TEZ/IVA vs. healthy
Cell activation involved in immune response
Leukocyte activation involved in immune response
Neutrophil activation involved in immune response
Neutrophil degranulation
Myeloid leukocyte activation
Myeloid cell activation involved in immune response
Leukocyte degranulation
Positive regulation of GTPase activity
Regulation of GTPase activity
Protein autophosphorylation
Reactive oxygen species metabolic process
Peptidyl−tyrosine phosphorylation
Peptidyl−tyrosine modification
Regulation of leukocyte mediated immunity
platelet activation
Lymphocyte activation
Positive regulation of immune effector process
Leukocyte differentiation
Lymphocyte proliferation
Myeloid leukocyte migration
Leukocyte migration
Regulation of reactive oxygen species metabolic process
Mononuclear cell proliferation
B cell activation
Immune system development
Regulation of leukocyte activation
Hematopoietic or lymphoid organ development
Chemotaxis
Taxis
Hemopoiesis
Positive regulation of cell adhesion
Regulation of cell activation
Positive regulation of response to external stimulus
Positive regulation of defense response
Positive regulation of cell activation
T cell activation
Positive regulation of immune system process
Positive regulation of kinase activity
Positive regulation of protein phosphorylation
Cytokine production
Regulation of MAPK cascade
Regulation of secretion by cell
Positive regulation of cell migration
Positive regulation of phosphorylation
Regulation of cytokine production
Cation transport
Regulation of immune system process
Positive regulation of phosphorus metabolic process
Positive regulation of phosphate metabolic process
Regulation of protein phosphorylation
NES
3
2
1
0
−1
−2
−3

b)

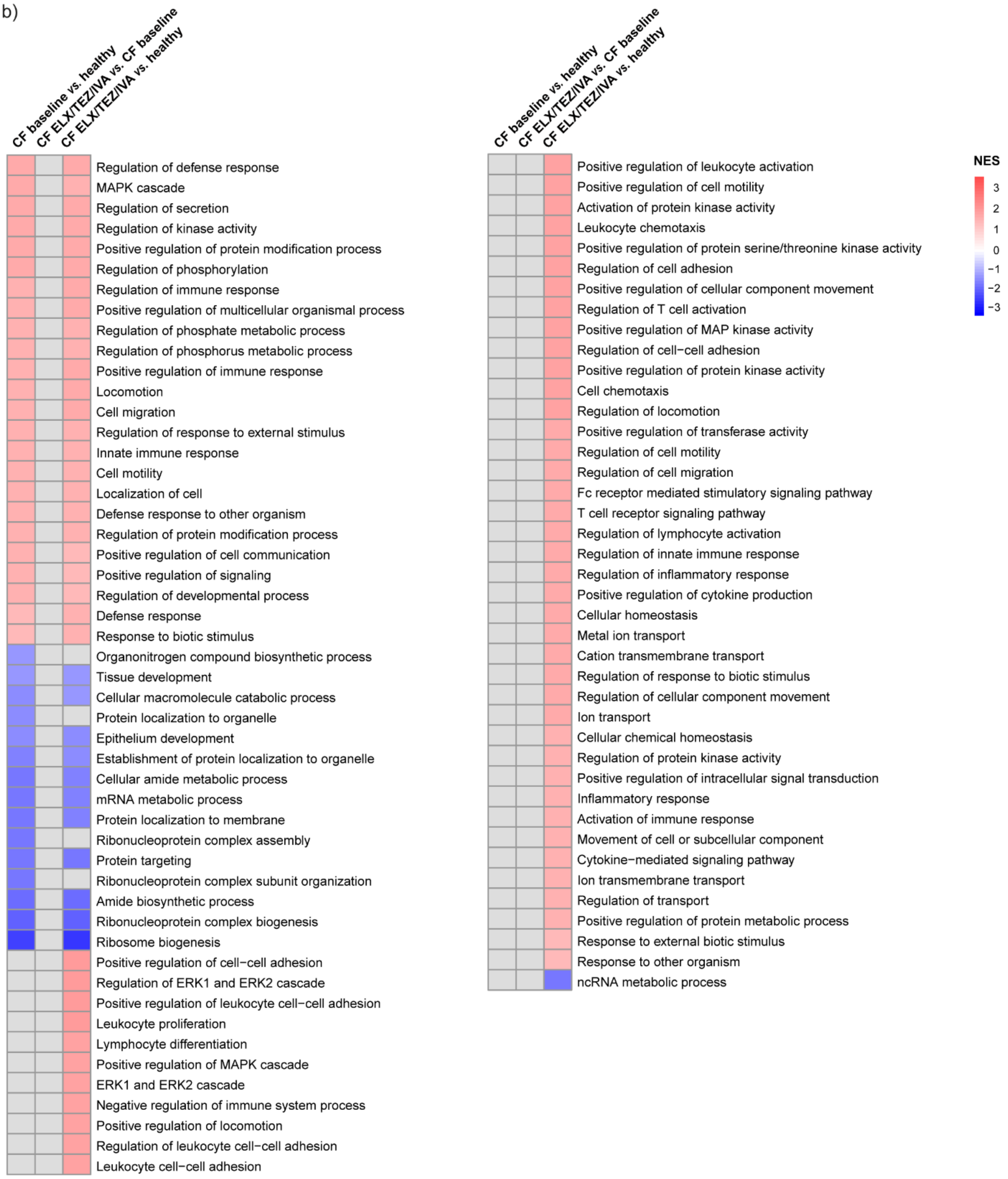


**Supplementary figure S3.** (a-b) Heatmaps showing enriched gene ontology (GO) terms of biological processes in sputum of patients with cystic fibrosis at baseline compared to healthy controls (first column), after initiation of elexacaftor/tezacaftor/ivacaftor (ELX/TEZ/IVA) compared to baseline (second column) and after initiation of ELX/TEZ/IVA compared to healthy controls (third column) ordered by normalized enrichment score (NES).
